# Brain Injury in COVID-19 is Associated with Autoinflammation and Autoimmunity

**DOI:** 10.1101/2021.12.03.21266112

**Authors:** EJ Needham, AL Ren, RJ Digby, JG Outtrim, DA Chatfield, AE Manktelow, VFJ Newcombe, R Doffinger, G Barcenas-Morales, C Fonseca, MJ Taussig, RM Burnstein, C Dunai, N Sithole, NJ Ashton, H Zetterberg, M Gisslen, A Edén, E Marklund, MJ Griffiths, J Cavanagh, G Breen, SR Irani, A Elmer, N Kingston, JR Bradley, LS Taams, BD Michael, ET Bullmore, KGC Smith, PA Lyons, AJC Coles, DK Menon, the Cambridge NeuroCOVID Group, the NIHR COVID-19 BioResource, Cambridge NIHR Clinical Research Facility

**Author notes:** **Cambridge NeuroCOVID Group:** Anwar F, Allinson K, Bhatti J, Bullmore ET, Chatfield DA, Christmas D, Coles AJ, Coles JP, Correia M, Das T, Fletcher PC, Jubb AW, Lupson VC, Manktelow AE, Menon DK, Michell A, Needham EJ, Newcombe VFJ, Outtrim JG, Pointon L, Rodgers CT, Rowe JB, Rua C, Sithole N, Spindler LRB, Stamatakis EA, Taylor J, Valerio F, Widmer B, Williams GB. **NIHR COVID-19 BioResource^22^:** Kingston N, Graves B, Le Gresley E, Caputo D, Stark H, Townsend P, Stirrups KE, Chinnery PF, Bradley JR. **NIHR Cambridge Clinical Research Facility:** Saunders C, Elmer A.

## Abstract

COVID-19 has been associated with many neurological complications including stroke, delirium and encephalitis. Furthermore, many individuals experience a protracted post-viral syndrome which is dominated by neuropsychiatric symptoms, and is seemingly unrelated to COVID-19 severity. The true frequency and underlying mechanisms of neurological injury are unknown, but exaggerated host inflammatory responses appear to be a key driver of severe COVID-19 more broadly.

We sought to investigate the dynamics of, and relationship between, serum markers of brain injury (neurofilament light [NfL], Glial Fibrillary Acidic Protein [GFAP] and total Tau) and markers of dysregulated host response including measures of autoinflammation (proinflammatory cytokines) and autoimmunity. Brain injury biomarkers were measured using the Quanterix Simoa HDx platform, cytokine profiling by Luminex (R&D) and autoantibodies by a custom protein microarray.

During hospitalisation, patients with COVID-19 demonstrated elevations of NfL and GFAP in a severity-dependant manner, and there was evidence of ongoing active brain injury at follow-up 4 months later. Raised NfL and GFAP were associated with both elevations of pro-inflammatory cytokines and the presence of autoantibodies; autoantibodies were commonly seen against lung surfactant proteins as well as brain proteins such as myelin associated glycoprotein, but reactivity was seen to a large number of different antigens.

Furthermore, a distinct process characterised by elevation of serum total Tau was seen in patients at follow-up, which appeared to be independent of initial disease severity and was not associated with dysregulated immune responses in the same manner as NfL and GFAP.

## Introduction

COVID-19 has been associated with several neurological complications including stroke and immune-mediated disorders such as Guillain-Barré syndrome and autoimmune encephalitis.^1^ Furthermore, up to a third of infected individuals experience a protracted post-viral syndrome following COVID-19 which is potentially of CNS origin given the dominance of neuropsychiatric symptoms such as fatigue and subjective cognitive difficulties.^2–4^ While the occurrence of physical brain injury is overt in some COVID-19-associated neurological syndromes such as stroke and encephalitis, a number of studies have suggested that brain injury can occur in the context of COVID-19 even in the absence of a clear concomitant neurological diagnosis. However, the mechanism that might drive this process requires further attention. ^5–15^ In COVID-19 disease, exaggerated host inflammatory responses appear to be a key driver of severe disease, and the most effective established therapies for systemic COVID-19 aim to attenuate this response.^16,17^ Initial attention focused on the innate immune system as a key driver, and emerging evidence also suggests a significant role for dysregulated adaptive immune responses.^18^ This combined maladaptive response is reminiscent of that seen in a spectrum of immune-mediated diseases – which extend from autoinflammatory to autoimmune in nature - described in non-COVID-19 settings.^19^

Here, we seek to investigate markers of a dysregulated immune host response, including surrogates of autoinflammation (proinflammatory cytokines) and autoimmunity (autoantibodies), and how they correlate with biomarkers of brain injury.

## Methods

### Study populations

Patients admitted to Cambridge University Hospital, UK with PCR-proven COVID-19 were identified between March 2020 and March 2021. Providing research personnel were available, all patients admitted to Cambridge were approached for consent, either in the acute phase, or at follow-up visit. The cohort of patients recruited from Cambridge were supplemented by a convenience sample of PCR-proven COVID-19 patients from Sahlgrenska University Hospital, Sweden (February – March 2020); previously included in a prospective sampling study.^20^ Written consent was gained from either patients themselves, or from their legal representatives where they lacked capacity to consent. Where written consent could not be gained due to restrictions on hospital visiting, legal representatives were consulted by telephone. This study was approved by the Swedish Ethical Review Authority (2020–01771) and the East of England – Cambridge Central Research Ethics Committee (17/EE/0025); via the Cambridge Biomedical Research Centre). Healthy controls were recruited through the Cambridge Biomedical Research Centre (prior to the COVID-19 pandemic) and all provided written consent (17/EE/0025). Data from a small positive control group consisting of patients with acute traumatic brain injury were included as a reference for the magnitude of brain injury biomarker elevations (REC 97/290).

### Procedures

Serum samples were collected at up to three timepoints from admission (acute [0-14 days], subacute [15–70 days] and convalescent [at outpatient follow up; >80 days). The samples were aliquoted, labelled with pseudoanonymised identifiers, and frozen immediately at −70°C. Samples from Sweden were then shipped on dry ice to the University of Cambridge.

### Demographic and clinical information

Demographic, clinical and laboratory information was recorded by the clinical team at the time of admission; Short Form Health Survey 36 (SF36)^21^ was completed in patients recruited to Cambridge University Hospital who returned for follow-up after their attendance to hospital. Patients were stratified into three groups of severity based on the treatment needed in the acute phase (Mild: no supplemental oxygen was required, Moderate: supplemental oxygen was required, Severe: invasive mechanical ventilation was required).

### Brain injury biomarker measurement

Neurofilament light, glial fibrillary acidic protein, total tau, and ubiquitin C-terminal hydrolase L1 concentrations were quantified in serum at the University of Cambridge using the Neurology 4-PLEX A assay run on an HD-X Analyser (Quanterix, Billerica, MA, USA). As per previous experience, UCH-L1 levels were predominantly around the lower level of quantification, with high coefficients of variance between replicates, and therefore were excluded from analysis. Five samples taken from patients within 3 days of severe traumatic brain injury were also assayed to provide a frame of reference for magnitude of changes seen.

### Protein microarray autoantibody profiling

Autoantibody screening was performed using a custom central nervous system protein microarray based on the HuProt™ (version 4.0) platform.^22,23^ The microarray was devised in collaboration with Cambridge Protein Arrays Ltd. (Cambridge, UK) and CDI laboratories (Puerto Rico) to detect autoantibodies predominantly directed against central nervous system antigens (n = 51), but also to a number of blood-brain barrier (n = 5) and other tissue-specific (n = 94, covering organ systems including lung, heart and coagulation) antigens, as well as spike and nucleocapsid antigens (full antigen list detailed in **Supplemental Figure 1**). The microarrays consist of a glass microscope slide with a thin nitrocellulose coating, printed with quadruplicate spots of recombinant yeast-expressed whole proteins. Each slide accommodates up to 12 individual serum samples. Samples from healthy controls and patients with COVID-19 were randomly distributed across the slides to mitigate against experimental variation.

The slides were blocked in 2% BSA/ 0.1% PBS-Tween overnight at 4°C, washed, and then incubated with 200 μl of 1:1000 diluted serum at room temperature for two hours. The slides were washed again, incubated at room temperature for two hours with fluorophore-conjugated goat anti-human IgM-μ chain-Alexa488 (Invitrogen, Carlsbad, CA, USA, Cat. No. A21215) and goat anti-human IgG-Fc-DyLight550 (Invitrogen Cat. No. SA5-10135) secondary antibodies, washed, and then scanned using a Tecan LS400 scanner and GenePix Pro v4 software, with the output being median fluorescence value of the quadruplicate spots for each protein.

### Cytokine Profiling

Serum concentrations of TNFα, IL-1β, IL-6, IL-10 and IFN-γ were quantified using by multiplexed particle based flow cytometry on a Luminex 200 analyser using xPonent Software (R&D Systems / Luminex) according to manufacturer’s recommendations. The population reference ranges derived for clinical use with this assay were utilised. Sensitivities / minimum detectable doses as indicated by the manufacturer are: IFN-γ (0.04 pg/ml); IL-1β (0.08 pg/ml); IL-6 (0.14 pg/ml); IL10 (0.21 pg/ml); TNFα (0.29 pg/ml).

### Statistical Analysis

Continuous descriptive data are presented using median and interquartile range, and categorical variables using number and percentage. Unpaired two-group comparisons were assessed using Mann-Whitney U tests, paired two-group comparisons with Wilcoxon Matched-Pairs Signed Rank tests and categorical comparisons with the Chi-squared statistic. Multiple t-tests were used to generate volcano-plots, with a false-discovery rate set to 1%. Comparisons between more than two groups were undertaken using Kruskal-Wallis test with post-hoc Dunn’s multiple comparison test. Correlations between continuous variables were assessed using Spearman’s rank correlation coefficient, and where multiple correlations were assessed within an experiment, Bonferroni correction was used to determine the appropriate level of significance. Principal component analysis was used as a dimension reduction technique to identify inflammatory cytokine profiles. All analyses were performed using GraphPad Prism Version 9.2.0.

### Protein microarray data analysis

As previously described,^23^ antibody binding was determined by measuring the median fluorescence intensity (MFI) of the four quadruplicate spots of each antigen; this value was then normalised by dividing it by the median MFI value of all antigens for that sample. These normalised values were then transformed into Z scores based on the distribution derived for each antigen from the healthy control cohort. A positive autoantibody “hit” was defined as an antigen where Z≥3.

## Results

### Study Populations

For brain injury biomarker analysis, 250 samples (from 175 patients; 122 from Cambridge and 53 from Gothenburg, at up to three time-points), and control samples from 59 age-matched healthy individuals were obtained. The 122 patients from Cambridge represented ∼7% of a total of 1666 patients admitted over the study period. Comparisons of the study population with the overall admitted population are shown in **Supplementary Figure 2**. Overall, there was no difference in age between patients and controls (51 [35-61] vs. 50 [32-62], but a larger proportion of males in the patient group (93 [53%] vs 21 [35%]; p = 0.02). Of the patients, 70 (40%) had mild disease, 72 (41%) moderate disease and 33 (19%) severe disease. The median (IQR) timings of the samples post-admission were: acute = 7 (3 – 10) days, subacute = 31 (26 – 35) days, and convalescent = 122 (109 – 136). A subset of these patients underwent autoantibody and cytokine profiling. Graphical description of these cohorts is shown in **Supplementary Fig. 2**.

### Neurofilament-light (NfL) and glial fibrillary acidic protein (GFAp) rise acutely in a severity-dependant manner, whilst elevated serum total tau concentrations are seen in the convalescent period irrespective of severity

In patients with COVID-19, serum concentrations of NfL and GFAP were raised in a severity-dependant manner at both the acute and subacute timepoints; there was no systematic difference between serum total tau concentrations between patients and controls (**Fig. 1A&B, Supplementary Table 1**).

**Figure 1.**
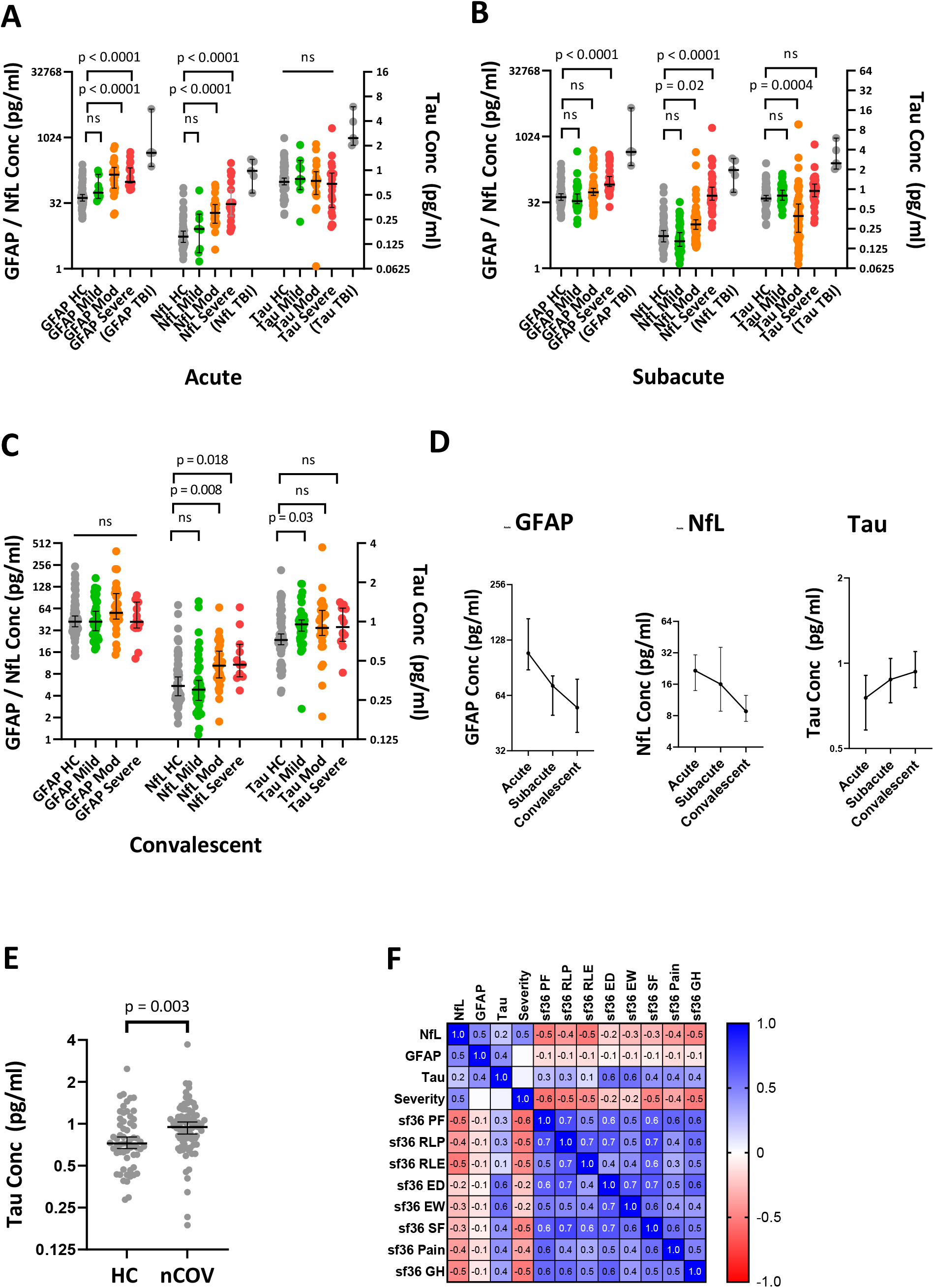
Serum brain injury biomarker concentrations in patients with COVID-19. **A-C**) Dotplots showing the effect of COVID-19 disease severity on brain injury biomarkers at the acute, subacute and convalescent timepoints; representative levels from five patients with acute severe traumatic brain injury (TBI) included as a reference for magnitude of elevation. **D**) Temporal changes in serum GFAP, NfL and Tau concentrations. **E**) Elevated serum total Tau concentrations at the convalescent timepoint in COVID-19. **F**) Correlation matrix of brain injury biomarkers and SF36 quality of life measure components *HC = healthy controls, nCOV = COVID-19, TBI = traumatic brain injury, CNS = central nervous system complication, PNS = peripheral nervous system complication. Multiple group comparisons are by Kruskal-Wallis test with post-hoc Dunn’s multiple comparison test; two-group unpaired comparisons are by Mann-Whitney U test, and paired by Wilcoxon matched-pairs signed rank test; correlations are by Spearman’s rank*.

The temporal dynamics, in 67 patients who provided longitudinal samples, showed that both GFAP and NfL tended to fall with time, although NfL rose in some patients between the acute and subacute timepoints, presumably as a result of its longer half-life (**Fig. 1D; Supplementary Fig. 3A**). Unusually, serum total tau concentrations were significantly higher than controls at the convalescent timepoint (0.95 [0.75 – 1.15] vs. 0.72 [0.60 – 1.04] pg / ml, p = 0.003; **Fig. 1D & E**).

At the convalescent timepoint, serum GFAP concentrations were no higher than controls irrespective of disease-severity, but serum NfL concentrations persisted at levels which were higher in patients who had developed moderate and severe COVID-19 compared with controls (**Fig. 1C, Supplementary Table 1**). The elevation of serum total tau concentration did not vary with severity, and indeed after correction for multiple comparisons only patients who had developed mild disease remained significantly higher than controls (**Fig. 1C, Supplementary Table 1**). Convalescent levels of both NfL and GFAP concentrations correlated with paired samples taken at the 15-42 day timepoint (ρ = 0.69, p = 0.0008 and ρ = 0.82, p < 0.0001 respectively), but total Tau did not (ρ = 0.27, p = 0.02), suggesting that the residual elevations of NfL and GFAP are reflective of events occurring during the acute illness, whereas the subsequent elevation of total Tau appears to be independent from any acute effects.

To explore the relationship between elevations of convalescent brain injury biomarkers and clinical outcomes, we studied correlations with the eight components of the SF-36. High serum NfL concentrations appeared to correlate most strongly with worse scores (notably: physical functioning [ρ = - 0.52, p = 0.03], general health [ρ = - 0.48, p = 0.05] and role functioning – emotional [ρ = - 0.53, p = 0.02]). The relationship between serum total tau concentrations and SF-36 domains, however, was very different, with higher concentrations seemingly associating with better scores, particularly in the emotional components (emotional wellbeing [ρ = 0.56, p = 0.02] and energy/ vitality [ρ = 0.56, p = 0.02]; **Fig. 1F**). None of the above comparisons withstood adjustments for multiple comparisons, however.

While the number of patients in this cohort with specific neurological syndromic diagnoses were small (mononeuritis multiplex n = 3, opsoclonus myoclonus n = 1, and peripheral neuropathy with concurrent encephalopathy n = 1), these patients appeared to have higher brain injury biomarker levels, with one patient showing biomarker levels an order of magnitude higher than other patients. However, numbers were too small to draw definitive inferences (**Supplementary Fig. 3B**).

### Autoantibodies against a wide range of tissues are seen in COVID-19, particularly in severe disease, and associate with a proinflammatory cytokine profile

The data were first assessed for any group-wise differences in reactivity to self-antigens between patients with COVID-19 and controls; volcano plots showed that not only did COVID-19 patients demonstrate clear IgG reactivity to SARS-CoV-2 spike protein and nucleocapsid, but also to surfactant protein A (SFTPA1), a lung surfactant protein, mutations of which result in pulmonary fibrosis (**Fig. 2A**).^24^ This increased reactivity was seen in both subacute and convalescent samples (**Fig. 2B**); reactivity to SFTPA1 in the subacute samples was stronger in patients with moderate and severe disease than in either those with mild disease or healthy controls (**Fig. 2C**). The presence of this autoantibody has not been previously described in COVID-19; furthermore, we have not detected it in cohorts of patients with traumatic brain injury (unpublished data), suggesting that it is not a common finding in critically ill patients more generally. No increased IgM reactivities were seen to any antigen in subacute COVID-19 samples compared with controls, but there was higher IgM reactivity to both spike protein and HLA-DRA in the convalescent samples.

**Figure 2.**
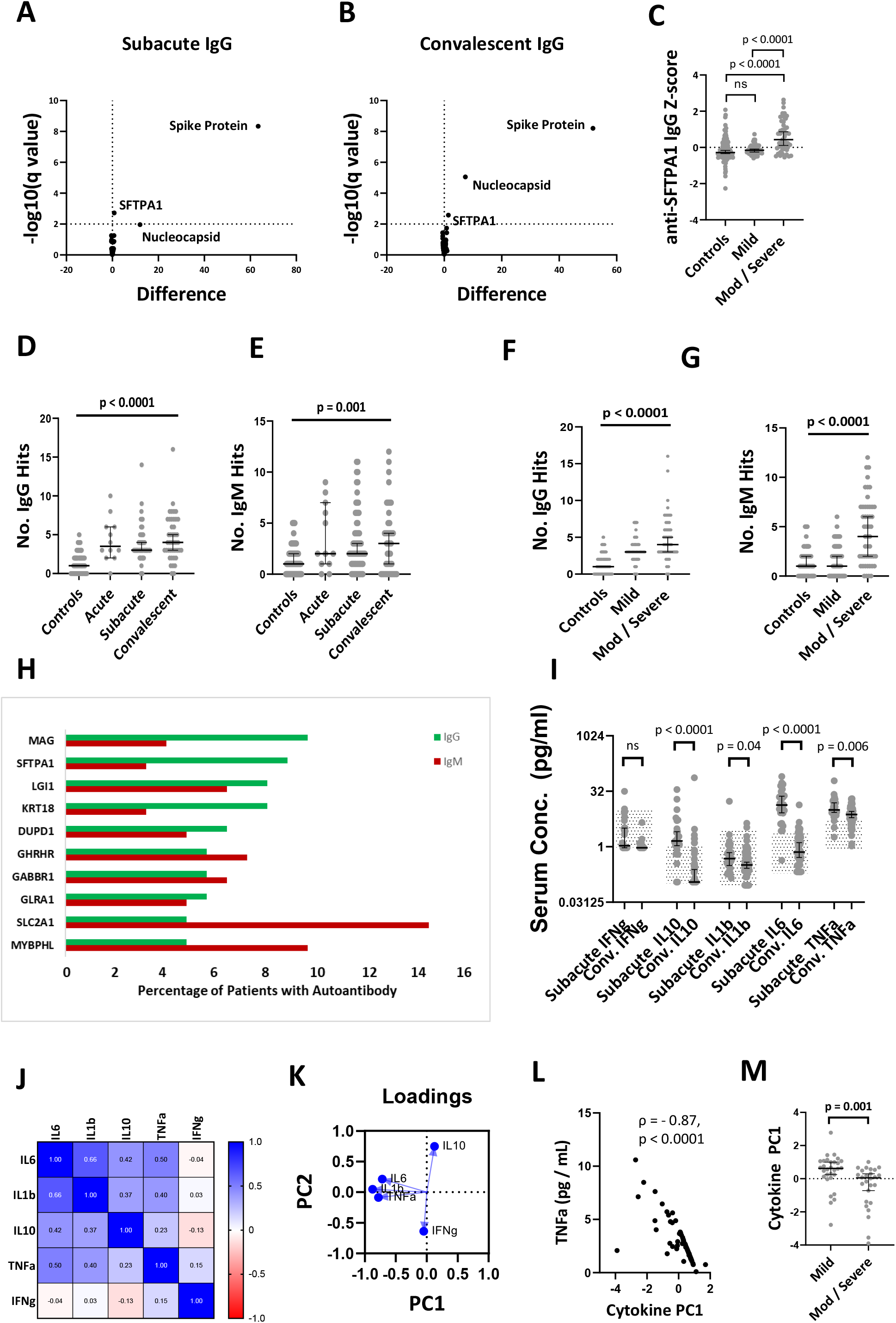
Autoantibody profiling in COVID-19. **A&B**) Volcano plots of groupwise comparisons in autoantibody profiles between COVID-19 patients and controls **C**) Relationship between disease severity and anti-SFTPA1 IgG autoantibodies **D&E**) Temporal profiles of IgG and IgM autoantibody responses **F&G**) Effect of disease severity on number of IgG and IgM autoantibody “hits”. **H**) Top ten most frequently detected autoantibodies across all samples. **I**) Comparison of cytokine profiles at the subacute and convalescent timepoints, with normal range shown by hatching. **J**) Correlation matrix between measured subacute cytokines **K**) Loadings plot from principal component analysis demonstrating the contributions of proinflammatory cytokines to PC1 **L)** Comparison in subacute proinflammatory cytokine response between mild and moderate / severe disease. *Volcano plots use multiple Mann-Whitney U tests with a false-discovery rate set to 1%; Multiple group comparisons are by Kruskal-Wallis test with post-hoc Dunn’s multiple comparison test; two-group unpaired comparisons are by Mann-Whitney U test, correlation matrix is by Spearman’s rank*

While the group level comparisons provided information about pervasive autoantibody responses that were common across patients, this approach was less useful in identifying autoantibody responses which were found in a minority of patients but were still biologically interesting. Autoantibody profiles of the groups were therefore compared by assessing the number and targets of positive autoantibody hits to specific target antigens. COVID-19 patients had higher numbers of both IgG and IgM autoantibody hits than healthy controls, which peaked at the subacute timepoint, but remained elevated in the convalescent samples (**Fig. 2D&E**). Patients with moderate or severe disease had higher numbers of autoantibody hits than those with mild disease at the subacute timepoint (**Fig. 2F&G**), and the number of IgM and IgG autoantibodies an individual had were related (ρ = 0.32, p = 0.01).

Autoantibodies to many different antigens were seen, but some were seen more frequently (**Fig. 2H**). Anti-myelin associated glycoprotein (MAG) was the most commonly detected IgG autoantibody, seen in 9.6% COVID-19 samples but not seen in any healthy controls, followed by surfactant protein A (SFTPA1), which was detected in 8.8% patients, and again not seen in healthy controls (**Frequency of positive autoantibody hits in control and COVID-19 cohorts shown in Supplementary Table 2**). No specifically characteristic autoantibody was seen in the five patients with syndromic neurological diagnoses.

Elevations in serum cytokine concentrations were seen in the subacute samples, particularly IL-6, TNFα and IL-10, but many patients demonstrated concentrations persisting above the normal range in the convalescent samples. (**Fig. 2I**). There was substantial covariance between all cytokines other than interferon gamma (**Fig. 2J**), but principal component analysis demonstrated the three canonical pro-inflammatory cytokines driving PC1 (**Fig. 2K**; note that the proinflammatory cytokines generate a negative eigenvector [**Fig. 2L**]). Patients with moderate and severe disease demonstrated higher concentrations of proinflammatory cytokines (**Fig. 2M**). The number of both IgG and IgM hits correlated with an elevated proinflammatory cytokine response (PC1 from principal component analysis vs. IgG: ρ = −0.33, p = 0.01, PC1 vs. IgM: ρ = −0.30, p = 0.02).

### Magnitude of autoantibody and pro-inflammatory cytokine response associates with high serum markers of brain injury

To understand whether there was a relationship between inflammatory profiles and brain injury biomarkers, we compared brain injury biomarker levels with cytokines and autoantibody responses. At the subacute timepoint, serum GFAP and NfL concentrations positively correlated with both the number of IgG hits and increased proinflammatory cytokine responses (GFAP and NfL vs. IgG hits: ρ = 0.26, p = 0.03 and ρ = 0.38, p = 0.001 respectively [**Fig. 3A&B**]; GFAP and NfL vs. cytokine PC1 ρ = - 0.53, p < 0.0001 and ρ = −0.65, p < 0.0001 respectively), but there was no such relationship between serum total Tau concentration and number of IgG hits or cytokine response (ρ = 0.02, p = 0.90 and ρ = 0.17, p = 0.2). The number of IgM hits also correlated with serum NfL concentration (ρ = 0.33, p = 0.006), but not with GFAP or total Tau (ρ = 0.20, p = 0.10, and ρ = 0.07, p = 0.57 respectively). The relationship between brain injury biomarkers and the top 10 most frequently detected autoantibodies was investigated; after Bonferroni correction, serum NfL concentrations were associated with the Z score of IgG autoantibodies against NfL, SFTPA1 and MYBPHL (ρ = 0.35, p = 0.002, ρ = 0.38, p = 0.001 and ρ = 0.41, p = 0.0005 respectively), but none of the top 10 autoantibodies retained significance against serum GFAP or total Tau concentrations after correcting for multiple comparisons.

**Figure 3.**
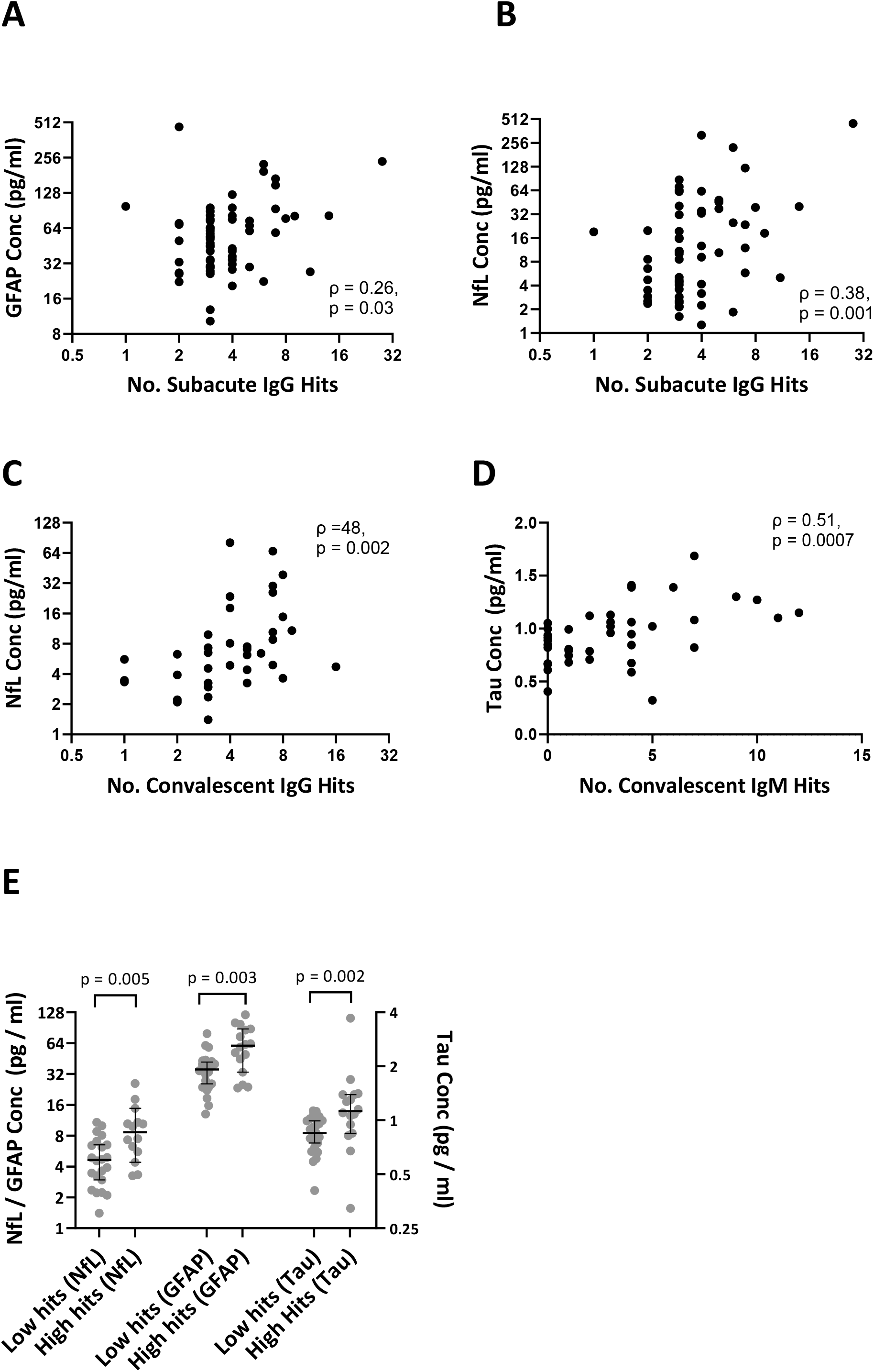
Relationship between serum brain injury biomarkers and autoantibody profiles. **A&B**) Correlation between number of IgG hits and serum GFAP and NfL concentrations at the subacute timepoint. **C**) Correlation between number of IgG hits and serum NfL concentrations at the convalescent timepoint. **D**) Correlation between number of IgM hits and serum total Tau concentrations at the convalescent timepoint. **E**) Comparison of convalescent serum brain injury biomarker concentrations between patients with high IgM responses (≥3 IgM hits Z≥3) versus those with low IgM responses (<3 IgM hits Z≥3). *Two-group unpaired comparisons are by Mann-Whitney U test, correlations are by Spearman’s rank*

In the convalescent period, the number of IgG hits once again correlated with serum NfL concentrations (ρ = 0.48, p = 0.002; **Fig. 3C**), but not GFAP or total tau (ρ = 0.12, p = 0.46, ρ = −0.08, p = 0.63 respectively). The relationship between brain injury biomarkers and cytokine profiles seen in the acute phase was replicated in convalescent patients, with elevations in proinflammatory cytokines associating with raised NfL and GFAP, but not total tau (PC1 vs. NfL: ρ = −0.55, p < 0.0001; GFAP: ρ = −0.26, p = 0.05; total Tau ρ = −0.1, p = 0.43).

### IgM autoantibodies in the convalescent phase are associated with elevation of brain injury biomarkers, especially Tau

At the convalescent timepoint, however, there was an association between number of IgM hits and all brain injury biomarkers, particularly total Tau (GFAP: ρ = 0.45, p = 0.004; NfL: ρ = 0.50, p = 0.001; total Tau: ρ = 0.51, p = 0.0007; **Fig. 3D**). To investigate this relationship further, patients were dichotomised into either high IgM responder (≥3 IgM hits) versus low IgM responder (<3 IgM Hits) groups, and the levels of brain-injury biomarkers compared. Serum concentrations of all three biomarkers were higher in the high IgM responder group, but again total Tau was the most highly significant difference (GFAP: 58.2 [32.6 - 87.05] vs. 37.8 [23.8 – 43.1], p = 0.03; NfL: 7.5 [5.2 – 16.5] vs. 4.6 [3.0 – 8.1], p = 0.026; total Tau: 1.1 [0.9 – 1.3] vs. 0.8 [0.7 – 0.9], p = 0.001; **Fig. 3E**).

## Discussion

The aim of this study was to examine how frequently brain injury occurred in COVID-19, both acutely and in convalescence, and whether elevated brain injury biomarkers were associated with a dysregulated host inflammatory response. We demonstrated that brain injury biomarkers are elevated in a severity-dependent manner in the acute phase, and that these elevations are associated with both raised pro-inflammatory cytokines and the presence of autoantibodies. When patients were followed up (∼four months post-admission), there was evidence that this immunological dysregulation had not fully resolved and was associated with serum markers of ongoing active brain injury (namely neurofilament light-chain), albeit to a lesser degree than in the acute illness. In addition, in convalescent patients, there appeared to be a second, separate, process, which was characterised by a different pattern of serum brain injury biomarkers (more specifically elevation of total Tau), which were not related to initial COVID-19 severity or pro-inflammatory cytokine levels but were associated with the presence of IgM autoantibodies. We observed autoantibody responses to many different targets (most commonly lung surfactant protein A1 and myelin associated glycoprotein), but the particular target of the autoantibody did not seem to relate to the presence of brain injury; rather, it seemed that the more diverse the autoantibody repertoire generated (reflecting a more generalised immune response), the more significant the degree of brain injury.

Our data confirms and extends previous studies investigating brain injury biomarkers in COVID-19, which have suggested that blood NfL concentrations are elevated in acute COVID-19 infection, and associate with severity of illness and therefore poor outcome.^5–15^ A longitudinal cohort study by members in our collaboration, demonstrated that serum NfL and GFAP levels had returned to baseline by six months following admission,^7^ suggesting that the persistent elevation in NfL at four months in our cohort is capturing the end of this period of active brain injury. The late elevations in total Tau seen in our cohort, however, are novel, as there is no precedent in the COVID-19 literature for this. Elevated serum total Tau concentrations have been described in patients with tauopathies such as Alzheimer’s Disease and Frontotemporal dementia,^25^ and serum concentrations associate with trajectory of cognitive decline in these conditions.^26,27^ Larger cohorts will be required to accurately delineate the association between late elevated total Tau and clinical outcome, however the lack of association between initial disease severity and subsequent total Tau elevation is tantalising: if replicated, this may represent an accessible biomarker to explore the basis of the protracted neuropsychological sequelae that occurs in a substantial minority of people infected with SARS-CoV-2.

It is well recognised that viral infections can trigger autoantibody production, both low-affinity polyreactive species, as well as higher-affinity specific species such as anti-cardiolipin antibodies.^28,29^ This phenomenon has been replicated in COVID-19, with a number of studies describing the presence of autoantibodies to a plethora of targets including “traditional” rheumatological autoantibodies as well as less clinically established autoantibodies such as those targeting type 1 interferons.^30–35^ The role of these autoantibodies is largely unknown. Although they appear to occur more commonly in severe illness, they may well simply represent an epiphenomenon of tissue damage (perhaps even a useful mechanism for debris clearance). However, it has been suggested that autoantibodies to certain targets (such as interferons) may predispose to severe disease,^36^ and it appears that immune-complex formation is a potent driver of secondary immune cell activation in COVID-19.^37^

The associations seen in our data between brain injury biomarkers and dysregulation of both innate and adaptive immune responses may represent autoinflammatory and autoimmune mechanisms that drive neurological injury. The well-documented impact of immune modulatory treatments in preventing severe COVID-19 provides strong evidence that a substantial component of the acute pathophysiology of COVID-19 relates to an unbridled and dysregulated host response, rather than damage caused directly by the virus. Our data suggest that brain injury occurring during acute COVID-19 may also result from similar mechanisms, and provide a plausible mechanistic basis for these manifestations, given the scant evidence to support direct viral invasion of the brain by SARS-CoV-2.^1^

Our data do not define causality between the immunological parameters and the presence of brain injury. In the acute phase, both may be influenced by additional factors that drive severe disease. Indeed, the immunological changes may be occurring in response to tissue injury, rather than causing it. However, given the growing evidence of the detrimental effects of excess inflammation in COVID-19 more broadly, it is plausible that the elevation of brain injury biomarkers is driven by a maladaptive host response.^38^ This may be the result of neuroinflammation *per se*,^39–42^ or inflammatory injury to the cerebrovascular bed, which subsequently results in microvascular ischaemic brain injury.^43–46^ Similar considerations may apply to the convalescent phase of illness, where the association of IgM autoantibodies with serum Tau could represent a persisting immunological dyscrasia driving brain injury. The relative specificity of Tau at this phase of the illness may represent tissue specificity of the process (Tau is a dendritic and axonal marker).

It should be noted that this study does not contain a disease control group consisting of patients with alternative respiratory infections, such as influenza, to determine to what degree the brain injury biomarkers and immunological changes are seen in other comparable conditions. A single small study suggested that patients with bacterial pneumonia displayed higher blood markers of brain injury than patients with COVID-19,^9^ and we hypothesis that the processes described in this paper are likely to be relevant to severe illness more broadly. This being the case, the lessons learnt from COVID-19 may serve to help mitigate against the neurological sequelae of severe illness in the future.^47^

In conclusion, we have demonstrated that markers of brain injury are associated with dysregulated immunological responses in COVID-19, and that there may be a separate late process irrespective of initial disease severity which is characterised by elevated serum total Tau concentrations and the presence of IgM autoantibodies.

## Data Availability

All data produced in the present study are available upon request to the authors

## Acknowledgements

We thank all the healthcare professionals who were involved in the care of the patients recruited to this study, and particularly thank all the patients who took part.

We thank NIHR BioResource volunteers for their participation, and gratefully acknowledge NIHR BioResource centres, NHS Trusts and staff for their contribution. We thank the National Institute for Health Research, NHS Blood and Transplant, and Health Data Research UK as part of the Digital Innovation Hub Programme. The views expressed are those of the author(s) and not necessarily those of the NHS, the NIHR or the Department of Health and Social Care.

These studies were supported largely by the NIHR Cambridge Biomedical Centre and by NIHR funding to the NIHR BioResource (RG94028 & RG85445).

B.D.M. is supported by grants from the Medical Research Council (MR/V007181/1 and MR/T028750/1) and Wellcome (ISSF201902/3)

EN, DKM and AC are supported by Brain Research UK.

HZ is a Wallenberg Scholar supported by grants from the Swedish Research Council (#2018-02532), the European Research Council (#681712), Swedish State Support for Clinical Research (#ALFGBG-720931), the Alzheimer Drug Discovery Foundation (ADDF), USA (#201809-2016862), the AD Strategic Fund and the Alzheimer’s Association (#ADSF-21-831376-C, #ADSF-21-831381-C and #ADSF-21-831377-C), the Olav Thon Foundation, the Erling-Persson Family Foundation, Stiftelsen för Gamla Tjänarinnor, Hjärnfonden, Sweden (#FO2019-0228), the European Union’s Horizon 2020 research and innovation programme under the Marie Skłodowska-Curie grant agreement No 860197 (MIRIADE), and the UK Dementia Research Institute at UCL.

MG is supported by the Swedish State Support for Clinical Research (ALFGBG-717531) and by grants from the SciLifeLab National COVID-19 Research Program, financed by the Knut and Alice Wallenberg Foundation (KAW 2020.0182 and 2020.0241).

GB is supported by a sabbatical grant from PASPA-DGAPA-UNAM, México.

SRI is supported by a Wellcome Trust Fellowship [104079/Z/14/Z], a Medical Research Council Fellowship [MR/V007173/1], BMA Research Grants-Vera Down grant (2013) and Margaret Temple (2017), Epilepsy Research UK (P1201), the Fulbright UK-US commission (MS-Society research award) and by the NIHR Oxford Biomedical Research Centre. The views expressed are those of the author(s) and not necessarily those of the NHS, the NIHR or the Department of Health. For the purpose of Open Access, the author has applied a CC BY public copyright licence to any Author Accepted Manuscript version arising from this submission.

VFJN is an Academy of Medical Sciences / The Health Foundation Clinician Scientist.

EB is supported by an NIHR Senior Investigator award

DKM is supported by an NIHR Senior Investigator Award and European Union 7th Framework program

We would like to thank Addenbrooke’s Charitable Trust and the NIHR Cambridge Biomedical Research Centre for their funding, and the NIHR Cambridge Clinical Research Facility outreach team for enrolment of patients.

The members of the NIHR COVID-19 BioResource are John Allison, Gisele Alvio, Ali Ansaripour, Sharon Baker, Stephen Baker, Laura Bergamaschi, Areti Bermperi, Ariana Betancourt, Heather Biggs, Sze-How Bong, Georgie Bower, John R. Bradley, Karen Brookes, Ashlea Bucke, Ben Bullman, Katherine Bunclark, Helen Butcher, Sarah Caddy, Jo Calder, Laura Caller, Laura Canna, Daniela Caputo, Matt Chandler, Yasmin Chaudhry, Patrick Chinnery, Debbie Clapham-Riley, Daniel Cooper, Chiara Cossetti, Cherry Crucusio, Isabel Cruz, Martin Curran, Jerome D. Coudert, Eckart M.D.D. De Bie, Rnalie De Jesus, Aloka De Sa, Anne-Maree Dean, Katie Dempsey, Eleanor Dewhurst, Giovanni di Stefano, Jason Domingo, Gordon Dougan, Benjamin J. Dunmore, Anne Elmer, Madeline Epping, Codie Fahey, Stuart Fawke, Theresa Feltwell, Christian Fernandez, Stewart Fuller, Anita Furlong, Iliana Georgana, Anne George, Nick Gleadall, Ian G Goodfellow, Stefan Gräf, Barbara Graves, Jennifer Gray, Richard Grenfell, Ravindra K. Gupta, Grant Hall, William Hamilton, Julie Harris, Sabine Hein, Christoph Hess, Sarah Hewitt, Andrew Hinch, Josh Hodgson, Myra Hosmillo, Elaine Holmes, Charlotte Houldcroft, Christopher Huang, Oisín Huhn, Kelvin Hunter, Tasmin Ivers, Aminu Jahun, Sarah Jackson, Isobel Jarvis, Emma Jones, Heather Jones, Sherly Jose, Maša Josipović, Mary Kasanicki, Jane Kennet, Fahad Khokhar, Yvonne King, Nathalie Kingston, Jenny Kourampa, Emma Le Gresley, Elisa Laurenti, Ekaterina Legchenko, Paul J. Lehner, Daniel Lewis, Emily Li, Rachel Linger, Paul A. Lyons, Michael Mackay, John C. Marioni, Jimmy Marsden, Jennifer Martin, Cecilia Matara, Nicholas J. Matheson, Caroline McMahon, Anne Meadows, Sarah Meloy, Vivien Mendoza, Luke Meredith, Nicole Mende, Federica Mescia, Alice Michael, Alexei Moulton, Rachel Michel, Lucy Mwaura, Francesca Muldoon, Francesca Nice, Criona O’Brien, Charmain Ocaya, Ciara O’Donnell, Georgina Okecha, Ommar Omarjee, Nigel Ovington, Willem H. Owehand, Sofia Papadia, Roxana Paraschiv, Surendra Parmar, Ciro Pascuale, Caroline Patterson, Christopher Penkett, Marlyn Perales, Marianne Perera, Isabel Phelan, Malte Pinckert, Linda Pointon, Petra Polgarova, Gary Polwarth, Nicole Pond, Jane Price, Venkatesh Ranganath, Cherry Publico, Rebecca Rastall, Carla Ribeiro, Nathan Richoz, Veronika Romashova, Sabrina Rossi, Jane Rowlands, Valentina Ruffolo, Jennifer Sambrook, Caroline Saunders, Natalia Savinykh Yarkoni, Katherine Schon, Mayurun Selvan, Rahul Sharma, Joy Shih, Kenneth G.C. Smith, Sarah Spencer, Luca Stefanucci, Hannah Stark, Jonathan Stephens, Kathleen E Stirrups, Mateusz Strezlecki, Charlotte Summers, Rachel Sutcliffe, James E.D. Thaventhiran, Tobias Tilly, Zhen Tong, Hugo Tordesillas, Carmen Treacy, Mark Toshner, Paul Townsend, Carmen Treacy, Lori Turner, Phoebe Vargas, Bensi Vergese, Julie von Ziegenweidt, Neil Walker, Laura Watson, Jennifer Webster, Michael P. Weekes, Nicola K. Wilson, Jennifer Wood, Jieniean Worsley, Marta Wylot, Anna Yakovleva, Cissy Yong and Julie-Anne Zerrudo.

---

**Supplementary Table 1.** Brain injury biomarker data. Values shown are median [IQR]. *HC = healthy controls (n = 59); Mild (n = 70), Mod (Moderate; n = 72), and Severe (n = 33) relates to severity of COVID-19*.

|  | Acute (n = 48) | Subacute (n = 123) | Convalescent (n = 79) |
| --- | --- | --- | --- |
| **GFAP HC** | 42.2 [32.2 - 68.6] | 42.2 [32.2 - 68.6] | 42.2 [32.2 - 68.6] |
| **GFAP Mild** | 55.3 [43.7-113] | 34.5 [27.5 - 70.5] | 42.15 [26.2 - 79.2] |
| **GFAP Mod** | 143.5 [64.6-259] | 54.3 [40.7 - 80.7] | 55.8 [44.9 - 104.0] |
| **GFAP Severe** | 98.1 [87.3 - 208.5] | 82.2 [64 - 165.9] | 41.9 [34.5 - 74.6] |
| **NfL HC** | 5.5 [3.6 - 10.4] | 5.5 [3.6 - 10.4] | 5.5 [3.6 - 10.4] |
| **NfL Mild** | 8.2 [2.6 - 15.9] | 4.2 [2.8-9.1] | 4.8 [3.0 - 10.0] |
| **NfL Mod** | 19.2 [11.1 - 30.7] | 10.1 [5.1 - 16.0] | 10.4 [6.9 - 18.0] |
| **NfL Severe** | 30.6 [14.3 - 81.3] | 45.6 [30.8-98.7] | 10.8 [7.4 - 19.4] |
| **Tau HC** | 0.72 [0.60 - 1.04] | 0.72 [0.60 - 1.04] | 0.72 [0.60 - 1.04] |
| **Tau Mild** | 0.78 [ 0.65 - 1.21] | 0.80 [0.66 - 1.03] | 0.95 [0.75 - 1.11] |
| **Tau Mod** | 0.74 [0.49 - 1.10] | 0.39 [0.18 - 0.73] | 0.90 [0.71 - 1.27] |
| **Tau Severe** | 0.68 [0.33 - 1.02] | 0.94 [0.68 - 1.24] | 0.91 [0.73 - 1.27] |

**Supplementary Figure 1.**
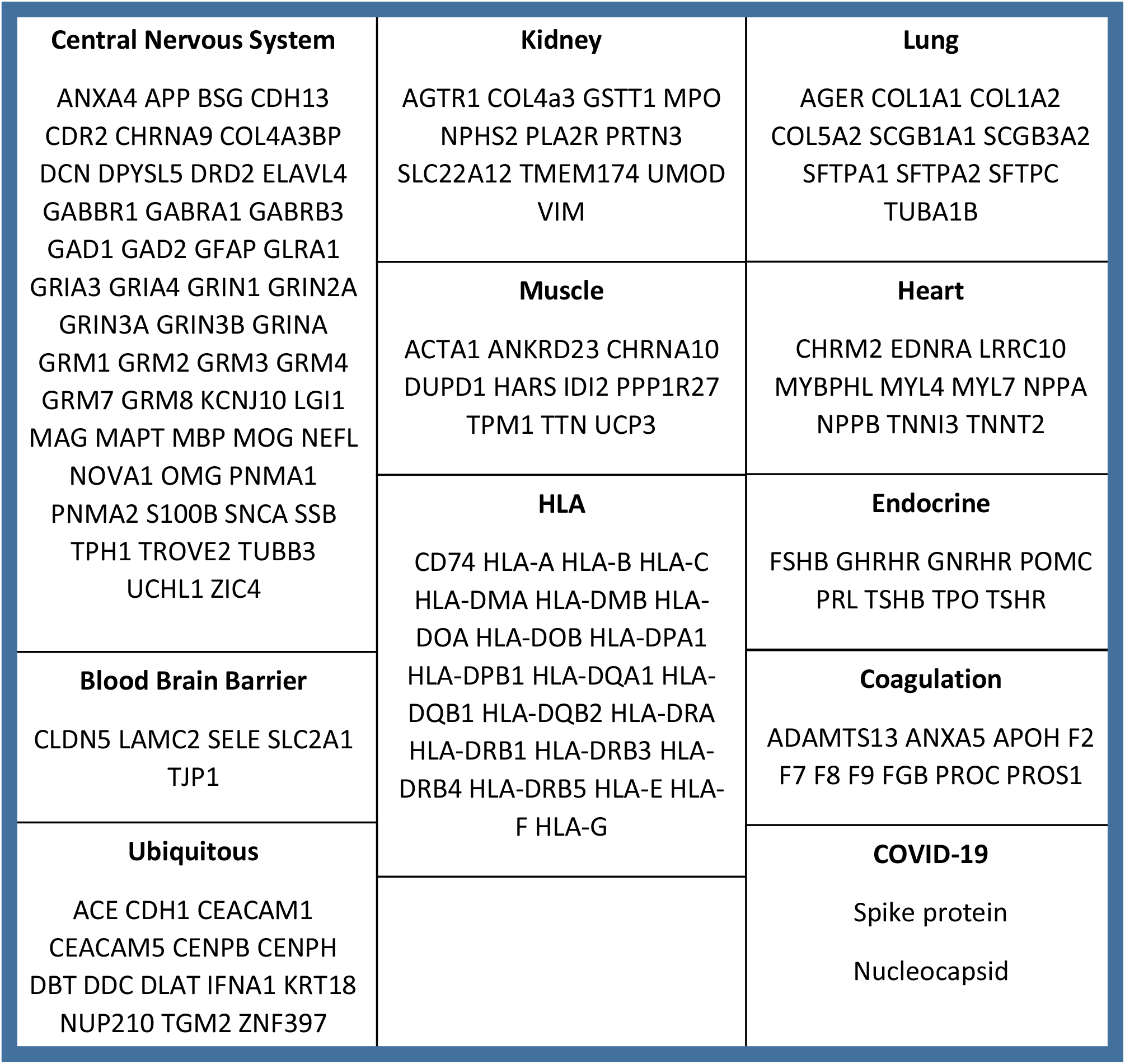
Protein microarray antigen composition

**Supplementary Figure 2.**
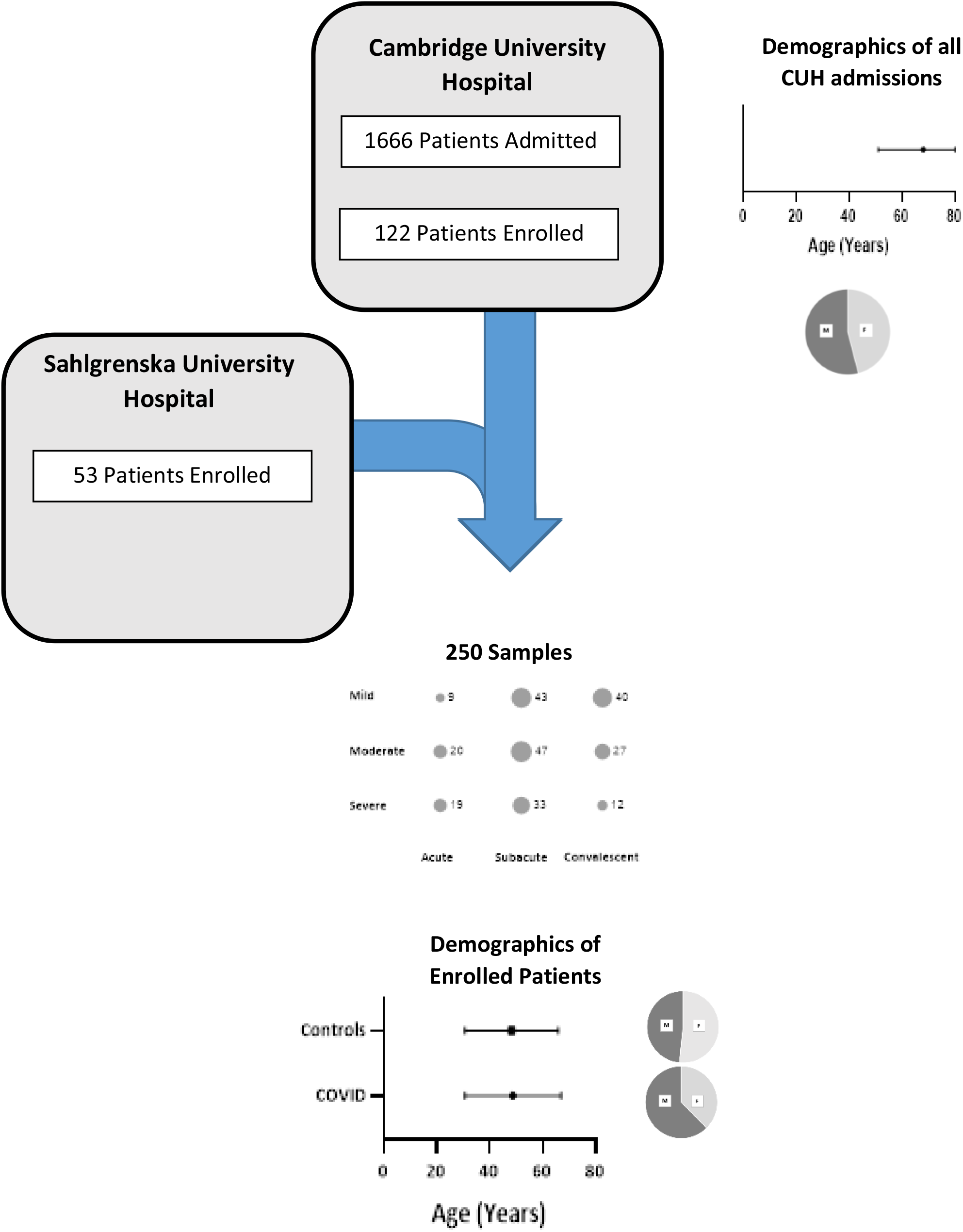
Demographic and sample details of patients and controls for both biomarker quantification and autoantibody profiling experiments.

**Supplemental Figure 3.**
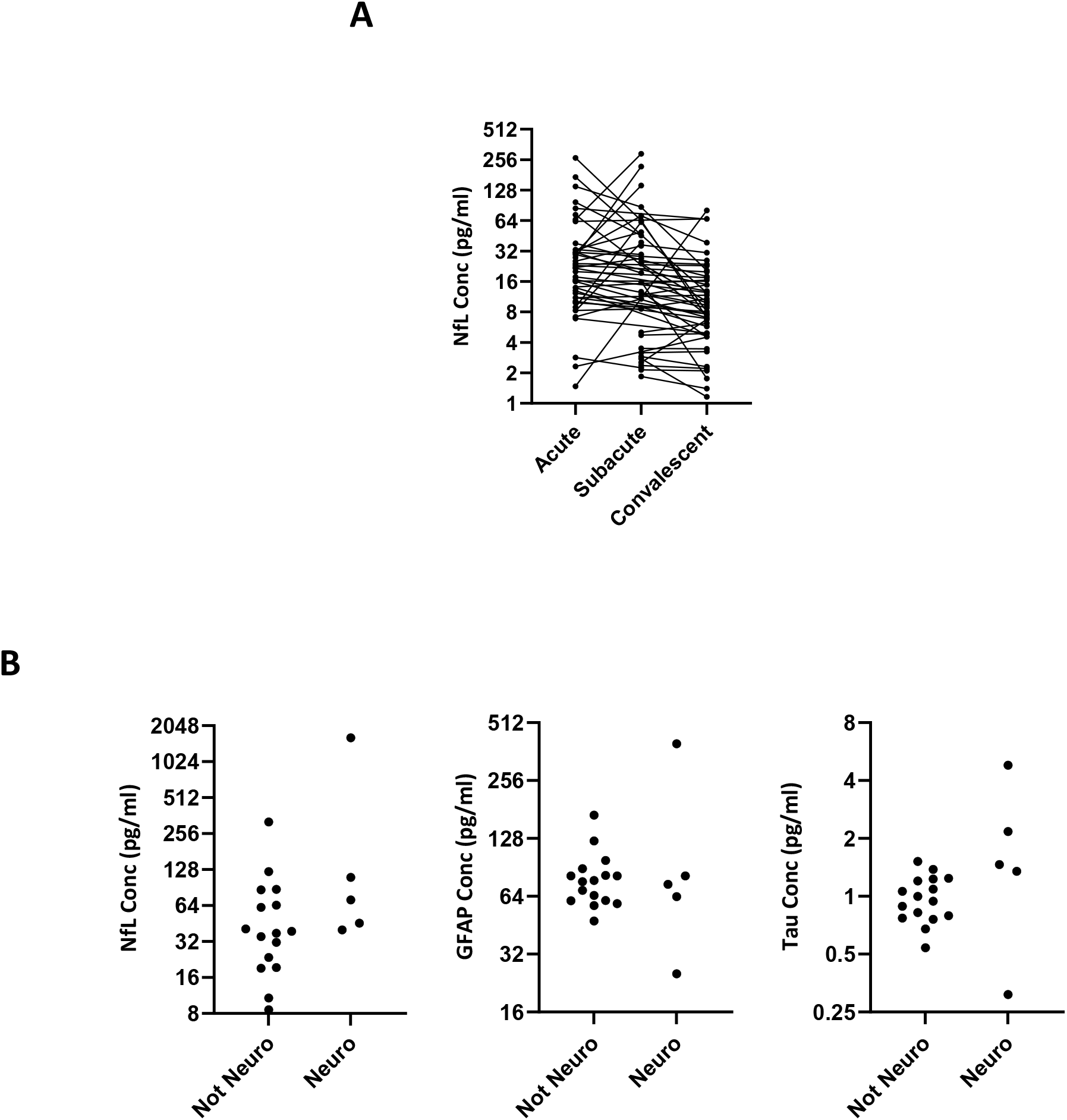
**A**) Spaghetti plot displaying temporal profiles of NfL in patients who contributed longitudinal samples **B**) Comparison of brain injury biomarkers between those patients with severe COIVD-19 who developed syndromic neurological diagnoses (mononeuritis multiplex n = 3, opsoclonus myoclonus n = 1, peripheral neuropathy with concurrent encephalopathy n = 1) versus those who did not.

**Supplementary Table 2.**
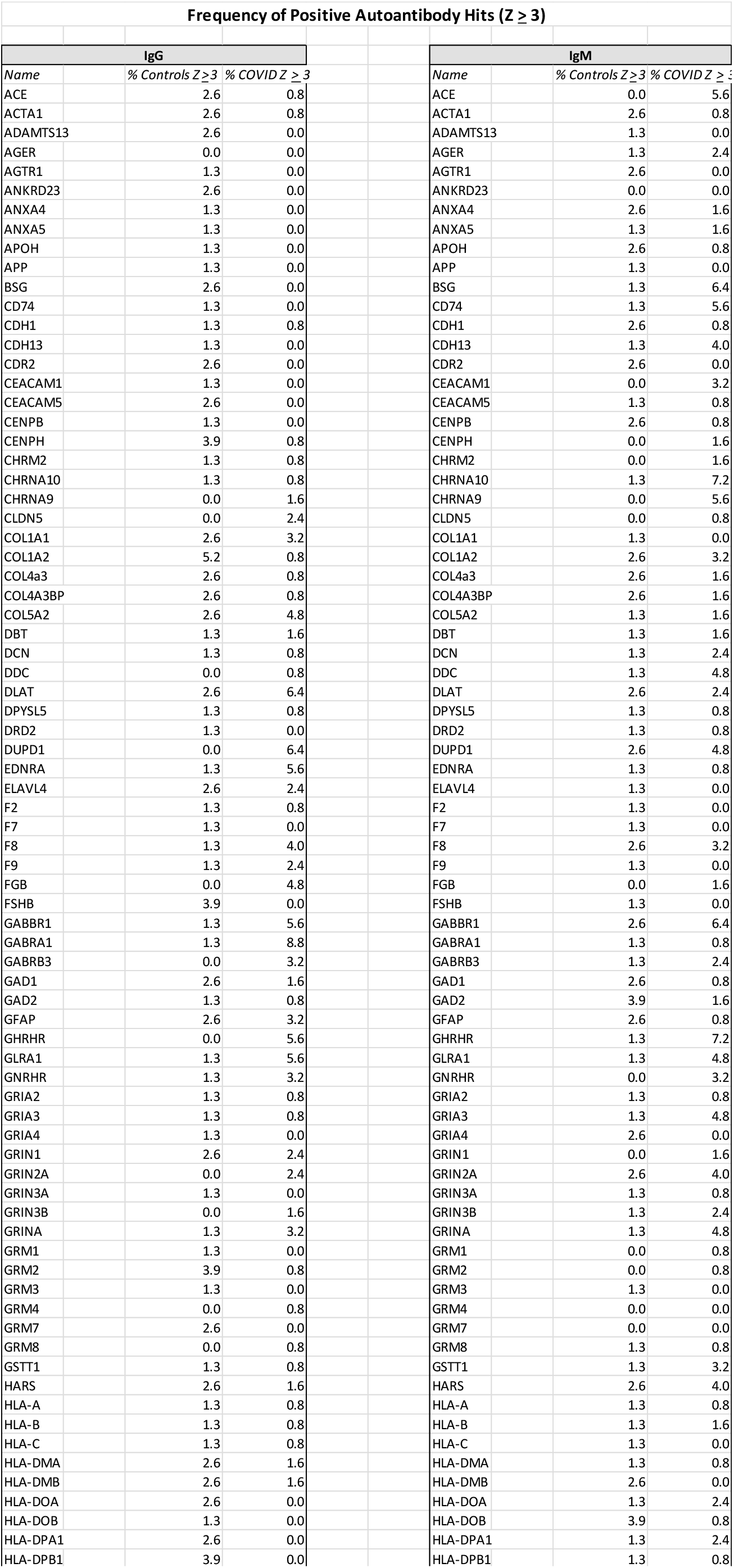
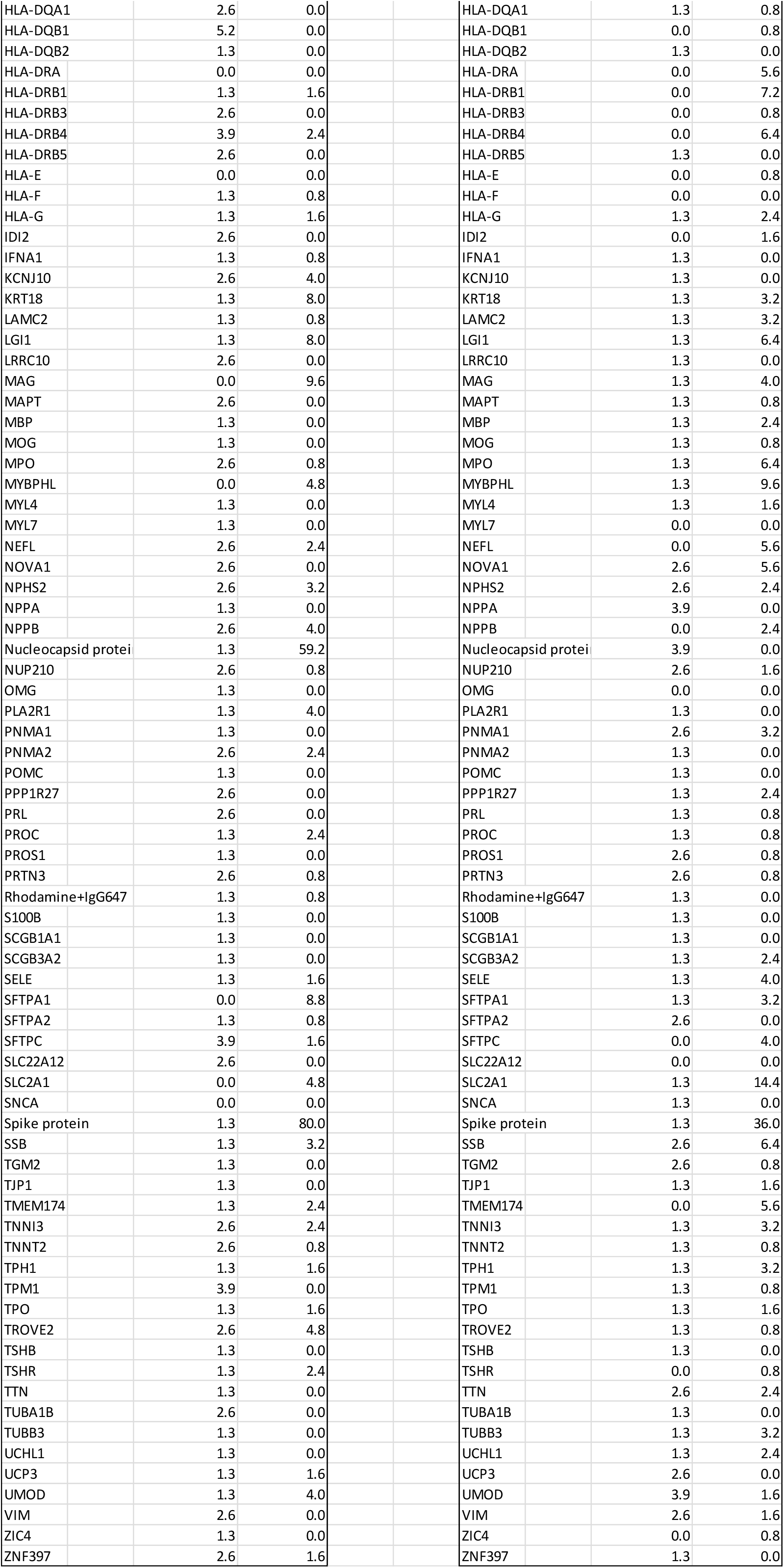
Frequency of Positive Autoantibody hits

